# In-Silo Federated Learning vs. Centralized Learning for Segmenting Acute and Chronic Ischemic Brain Lesions

**DOI:** 10.1101/2024.05.24.24307154

**Authors:** Joon Kim, Hoyeon Lee, Jonghyeok Park, Sang Hyun Park, Myungjae Lee, Leonard Sunwoo, Chi Kyung Kim, Beom Joon Kim, Wi-Sun Ryu

## Abstract

**Purpose:** To investigate the efficacy of federated learning (FL) compared to industry-level centralized learning (CL) for segmenting acute infarct and white matter hyperintensity.

**Materials and Methods:** This retrospective study included 13,546 diffusion-weighted images (DWI) from 10 hospitals and 8,421 fluid-attenuated inversion recovery images (FLAIR) from 9 hospitals for acute (Task I) and chronic (Task II) lesion segmentation. The mean ages (SD) for the training datasets were 68.1 (12.8) for Task I and 67.4 (13.0) for Task II. The frequency of male participants was 51.5% and 60.4%, respectively. We trained with datasets from 9 and 3 institutions for Task I and Task II, respectively, and externally tested them in datasets from 1 and 9 institutions each. For FL, the central server aggregated training results every four rounds with FedYogi (Task I) and FedAvg (Task II). A batch clipping strategy was tested for the FL models. Performances were evaluated with the Dice similarity coefficient (DSC).

**Results:** In Task I, the FL model employing batch clipping trained for 360 epochs achieved a DSC of 0.754±0.183, surpassing an equivalent CL model (DSC 0.691±0.229; p<0.001) and comparable to the best-performing CL model at 940 epochs (DSC 0.755±0.207; p=0.701). In Task II, no significant differences were observed amongst FL model with clipping, without clipping, and CL model after 48 epochs (DSCs of 0.761±0.299, 0.751±0.304, 0.744±0.304). Few-shot FL showed significantly lower performance. Task II reduced training times with batch clipping (3.5 to 1.75 hours).

**Conclusion:** Comparisons between CL and FL in identical settings suggest the feasibility of FL for medical image segmentation.

## Introduction

Numerous studies have investigated machine learning (ML) for segmenting medical images, demonstrating promising performance (1–3). Typically, these studies depend on preprocessed and aggregated open-source medical images. Injecting these algorithms into clinical practice is limited, however, partly due to suboptimal performance and domain shift (4). To address the representation gaps in open datasets, it is crucial to access diverse datasets from a broader range of imaging vendors and institutions. Unfortunately, legal constraints pose significant challenges for institutions and hospitals in sharing images with third-party servers for model training.

Unlike ML methodologies that depend on centralized data for training, federated learning (FL) (5,6) aggregates outcomes from individually trained models across multiple data silos. Upon receiving the model, clients train their local models with their private data. Upon completion, clients send their local model parameters back to the central server. The server then aggregates these submissions via an aggregation method to construct the next generation of the central model, which is sent it back to the clients for performance evaluation and declaration of a new round. By design, FL allows hospitals, acting as clients, to retain full privacy of their data while facilitating parallel, collaborative training across all partitioned datasets. With such benefits, FL emerged as a compelling alternative to centralized learning (CL) in applicative fields including of medical imaging.

Despite its advantages, FL is often underestimated due to concerns about potential underperformance compared to CL. One of the most significant limitations of FL is the potential of non-independent and identically distributed (non-i.i.d.) data, causing representation gaps between each client and divergence when aggregating (7). In practice, clients may have diverse distributions of images, including variations in signal-to-noise ratio, brightness, and disease prevalence. Furthermore, the requirement for constant communication with the central server in FL can pose challenging for certain medical institutions. Although one-shot or few-shot FL is under research (8), this approach has not been fully validated.

This study compared the efficacy of two cross-silo image segmentation FL models with industry-level CL models under similar conditions, employing a large-sample sized pragmatic non-i.i.d. brain MRI dataset. We compared FL models employing constant communication with those employing one-shot aggregation. Additionally, we introduce a “batch clipping” technique applied in FL training that accelerates the FL processes with negligible performance trade-off.

## Materials and Methods

Two datasets are used in this retrospective research: the acute ischemic lesion dataset (Task I) and the chronic ischemic lesion dataset (Task II). The datasets were used in previous works concerning the developments of the two CL models introduced in this paper. To compare the performance of a commercially used CL-based algorithm with an FL-trained algorithm, we adopted the CL architecture of the commercial software and a corresponding FL architecture for this comparison. Written informed consent was provided by patients or their legal representatives. The study protocol was approved by institutional review board of Dongguk University Ilsan Hospital (2017-09-017).

### Dataset

Details on datasets were available in Supplementary Material.

#### Acute ischemic lesion dataset

We consecutively enrolled 14,740 patients with ischemic stroke from 10 participating clients (Figure 1). After excluding 1,194 patients, leaving 13,546 patients amongst 9 institutions for training and validation and 1 institution for external test dataset. Previous study for CL utilized a total of 13,597 patients originated from 10 institutions, along with the same institution being used for the external test.

**Figure 1.**
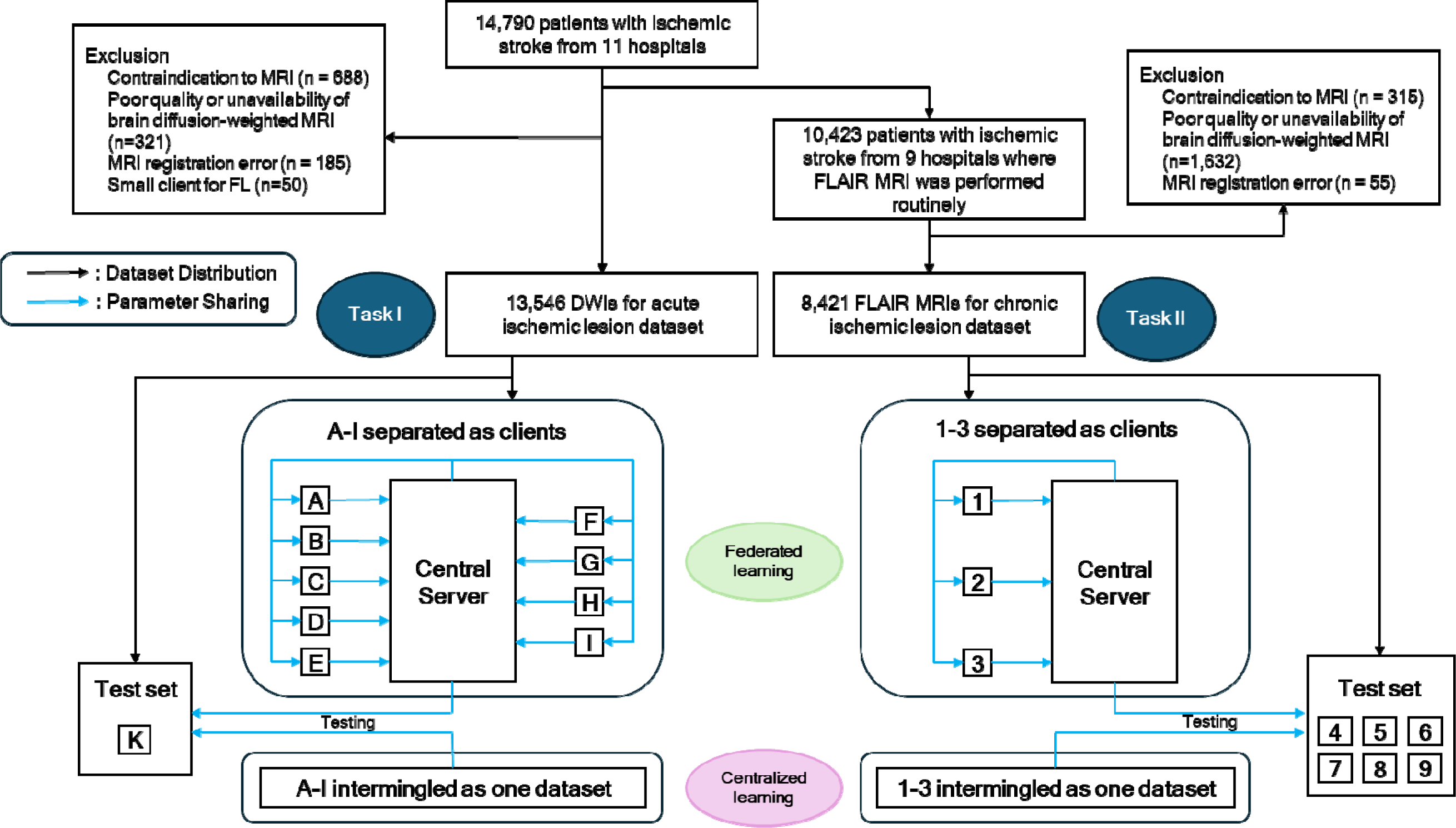
Study flow chart and study scheme for Task I, II. Blue arrows indicate the flow of model parameters, disseminated from the central server, trained by each client, and submitted to the server again. Exact distribution of the data can be found in the supplementary materials.

#### Chronic ischemic lesion dataset

We consecutively enrolled 10,423 patients admitted to 9 participating centers. After excluding 2,002 patients, leaving 8,421 patients amongst 3 institutions for training and 6 institutions for external test datasets. All data were forwarded from the previous CL study.

### Batch Clipping

The largest client possessed three times more data than the smallest, leading to inefficiencies in training, where small clients were kept idle for extended periods, awaiting one or two large clients. To reduce the total training time, we adopted a “batch clipping” strategy that caps the maximum number of batches per epoch. Even if certain data may not appear in one epoch, the expectation is that through the random batch shuffling over sufficient iterations, most data points will eventually be included in the training process. We constrained the number of batches to 200 and 1250 for Task I and II respectively (Figure 2), which corresponds to approximately 50∼55% of the largest client’s batch count. Clients with fewer batches than the clipping threshold remain unaffected, utilizing their entire dataset in every epoch. We experimented both tasks with and without batch clipping to investigate its effect.

**Figure 2.**
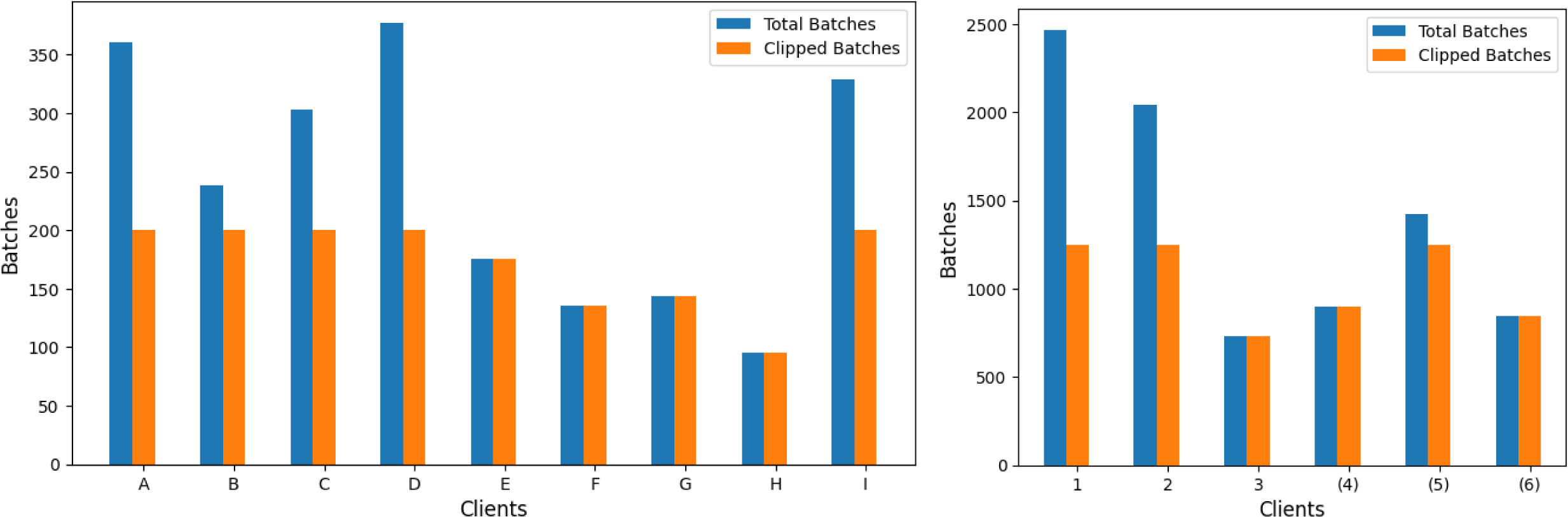
The Number of Total and Clipped Batches for Task I and II. The blue bar shows the number of batches of the entire dataset per client, and the red bar shows the clipped number of batches. Clipped clients stop training each epoch once it trains up to the clipping value. Clipping values are 200 for Task I and 1250 for Task B. Clients possessing less batches than the clipping value are unaffected.

### Implementation Details

FL models in both tasks are compared with an existing CL model with industry-level performance. The choice of ML model, number of epochs, hyperparameters, optimizers, and loss functions are matched as closely as possible with previous studies (Details in Supplementary Materials). To minimize communication overhead between server and clients, we implemented four epochs per round, indicating aggregation of results after 4 epoch of training in each client. We conducted both experiments using three RTX A6000 GPUs, with Flower (9) as the common FL framework.

We employed two aggregation methods in our study: FedAvg (5) and FedYogi (10). FedAvg is a straightforward method that averages the values of all received models on a coordinate-by-coordinate basis. FedYogi employs the Yogi (11) optimizer on the server side for more efficient updates and represents a more advanced variant of FedAvg (12).

#### Task I

segmented the acute ischemic lesion on diffusion-weighted image (DWI) using STU-Net (13). We partitioned the dataset into training and validation sets with a random sampling of a 4:1 ratio. The external test dataset for Task I consisted of 2777 DWIs acquired from a single institution not included in the training dataset. As the CL model for Task I utilized 1000 epochs for full convergence, we elected FedYogi for the aggregation method, which was expected to perform better than FedAvg over an elongated training. Both FL and CL models use PyTorch (14) as the ML framework. When evaluating the test Dice similarity coefficient (DSCs) over multiple rounds, we selected the first 100 DWIs of the external test dataset due to time constraints. Final DSC results were calculated with the entire external test dataset.

#### Task II

segmented the chronic ischemic lesion, known as white matter hyperintensities in FLAIR MRI, using U-Net (15). Task II dataset used the identical partition as the CL model, a 3:1:1 ratio for training, validation, and internal testing. However, we did not use the internal testing dataset for our experiments but rather used the external test datasets from another six institutions, ranging from 8422∼53344 slices per institution. In the Task II, we elected FedAvg as the aggregation method to show FL convergence without extensive hyperparameter tuning. Both FL and CL models use Tensorflow (16) as the ML framework. When evaluating the test DSCs over epochs 2∼12, we selected only the first 2000 slices of the test set due to time constraints. The final DSC results were calculated with the entire test set.

An additional study on Task II repositioned three institutions from the external test dataset to clients for training to prove the robustness of FL with varying clients. The three parenthesized clients in Figure 1 are the three clients excluded from the primary FL test results but involved in the additional study. All settings are held identical to the original Task II experiments.

### Statistical Analysis

To compare demographic and imaging characteristics between training and external test datasets, we used t-test, rank-sum test, AVONA, and Kruskal Wallis for continuous variables and chi-square test for categorical variables as appropriate.

To test the model performance in Task I and II, we used the DSC. All DSCs shown in the tables are averages and standard deviations of the dataset specified. For analyzing the main results, we employed paired sample t-tests per client to compare FL and CL models. In Task II, we utilized one-way ANOVA tests to compare DSC in each client in the external test dataset. For both statistical analyses, the p-value for significance is defined as less than 0.05. All calculations were performed using the scipy.stats(==1.10.1) library in Python(version 3.8.10, The Python Software Foundation).

## Results

### Patient Demographics

For Task I, the mean (SD) ages for the training&validation, and external test datasets were 68.1 (12.8) and 68.2 (12.4), respectively. There were significant differences in demographic and imaging characteristics between the training and external test datasets (Table S1). For Task II, the mean (SD) age for the training&validation was 67.4 (13.0) and 60.4% were men (Table S2). Mean ages for external test datasets ranged from 67.5 to 70.0. All variables significantly differed across the datasets, indicating dataset heterogeneity.

### Main Test Results

For Task I, the FedYogi FL models, with or without batch clipping, trained for 90 rounds (equivalent to 360 epochs) achieved a DSC of 0.754±0.183 and 0.762±0.173, respectively (Table 1). These results are better than those of the CL model trained for 360 epochs (DSC 0.691±0.229; p<0.001) and are comparable to the CL model trained for 940 epochs (DSC 0.755±0.207; p=0.701 and p=0.001), which is the best-performing CL model out of 1000 epochs. In turn, the 250-round trained FL models, with or without batch clipping exhibited a DSC of 0.776±0.170 and 0.776±0.169, respectively, outperforming the 940-epoch CL model (p<0.001). For Task II, segmenting chronic ischemic lesions, the FL Clipping, FL No Clipping, and CL models all performed similarly after 48 epochs (0.751±0.304, 0.761±0.299, 0.744±0.304).

**Table 1.**
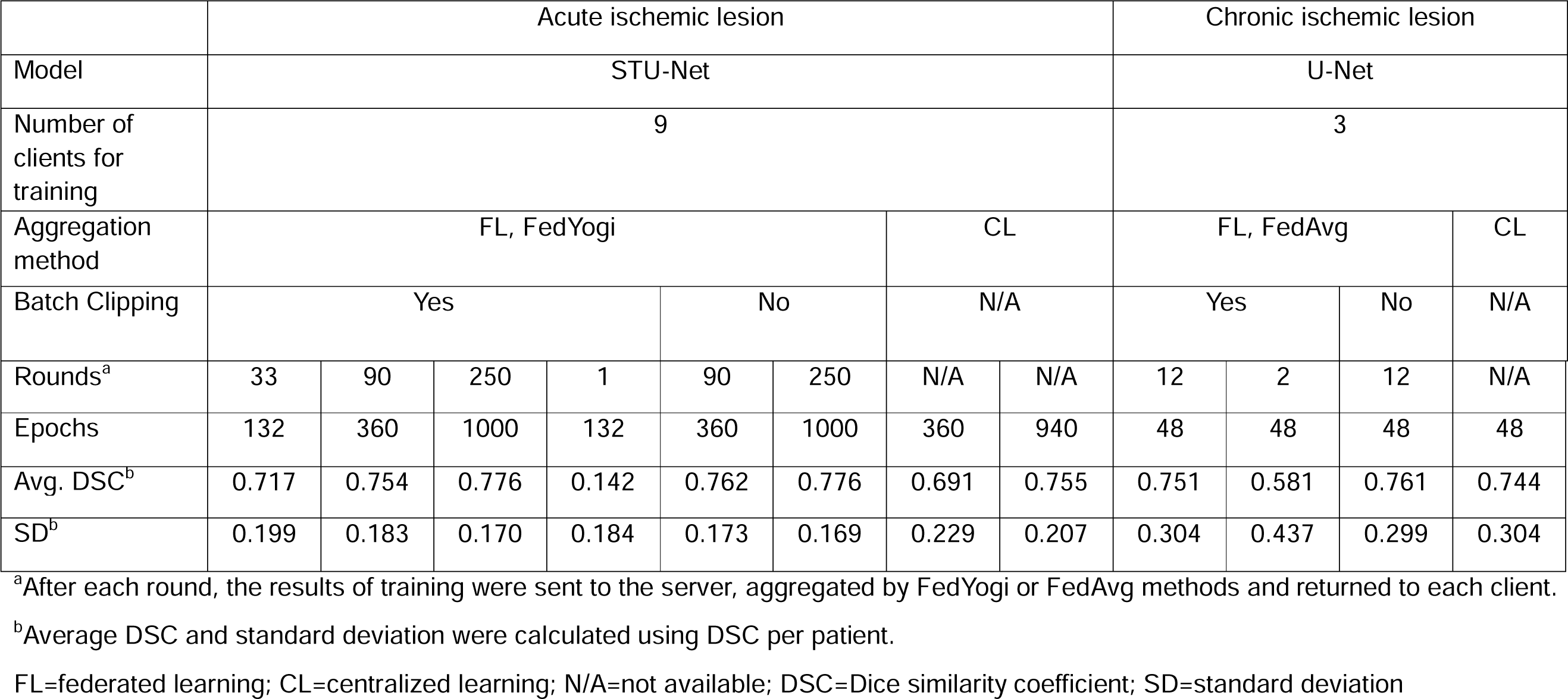
Comparison of segmenting performance between federated and centralized learnings.

For comparison, we also conducted few-shot FL experiments that aggregate only once or twice at the end of extensive decentralized learning. For Task I, a single round of FedYogi aggregation after training 132 epochs was not trained properly (DSC 0.142±0.184; p < 0.001 compared with FL model with 37 rounds). Task II tested a two-round, 24 epochs per round FedAvg FL, for an equivalent total epoch of 48. The result is significantly lower than FL with four epochs per round (DSC DSC 0.581±0.437; p < 0.001).

### Task I Convergence

Figures 3(a) and 3(b) depict training and validation losses for Task I. The CL model and all clipped FL clients properly converge while training, albeit at different values. The training and validation losses of FL without batch clipping is included in Figure S1. By the end of 1000 epochs, the CL model reaches 0.121 training loss (Figure 3(a)), while FL clients range from 0.113 to 0.182 in each training client with a weighted average of 0.144. For the validation loss (Figure 3(b)), the CL model reaches 0.126, whereas FL clients range from 0.098 to 0.176 with a weighted average of 0.138. Figures 3(c) compares the Test DSC for FL and CL models throughout their training and includes a smoothed-out graph for convenience. While the average FL losses were consistently higher than CL losses, FL consistently outperforms CL after 200 epochs in the test dataset, converging to a higher DSC by the end of the training.

**Figure 3.**
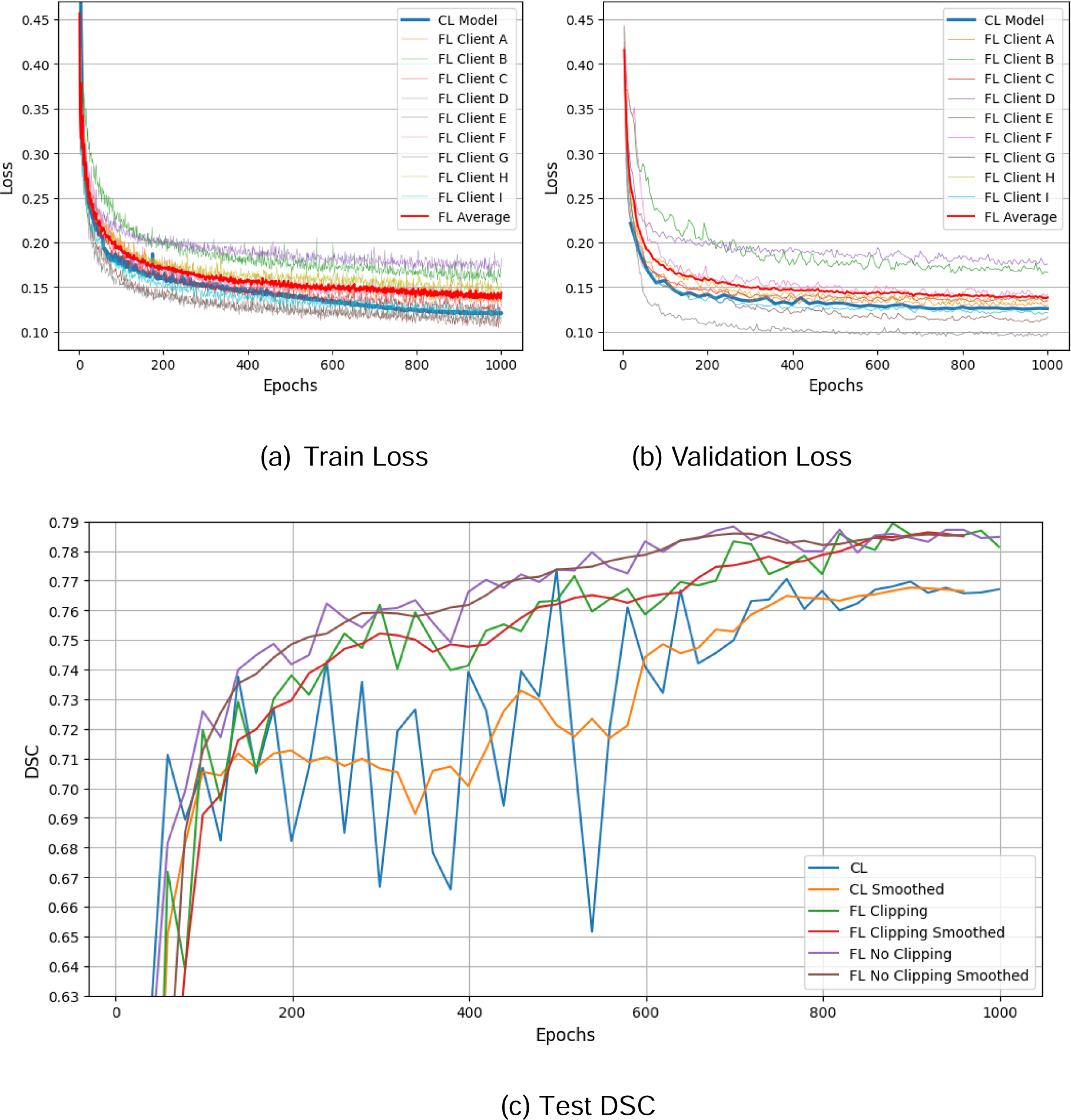
STU-Net Task I Losses and Test DSCs. a) Training loss of the CL model and clipped FL clients 0-8, up to 1000 epochs. b) Validation Loss of the CL model and clipped FL clients 0-8, up to 1000 epochs. Validation was performed every 4 epochs after aggregation. c) DSC of the CL model and aggregated FL models evaluated with the first 100 test set images, up to 1000 epochs. A smoothed out version using convolution with 5 previous results are drawn for ease of comparison.

### Task II Convergence

Figures 4(a) and 4(b) show the validation DSC for each FL training client in Task II, with and without batch clipping. The test DSC scores are lower than the validation DSC as the test set originates from new clients and possibly has a different distribution. Figure 4(c) compares the test DSC of the Clipping, No Clipping, and CL models. All three models converge to ∼0.75 DSC by the end of 48 epochs (equivalent to 12 rounds). Table 2 summarizes the average test DSC per each client in the external test dataset. While there are disparities amongst test clients, a one-way ANOVA test with the average DSC results reveals that their variances are not statistically significant (p=0.650). The FL model without clipping took around 3.5 hours to train 48 epochs, whereas FL with clipping took only about 1.75 hours.

**Figure 4.**
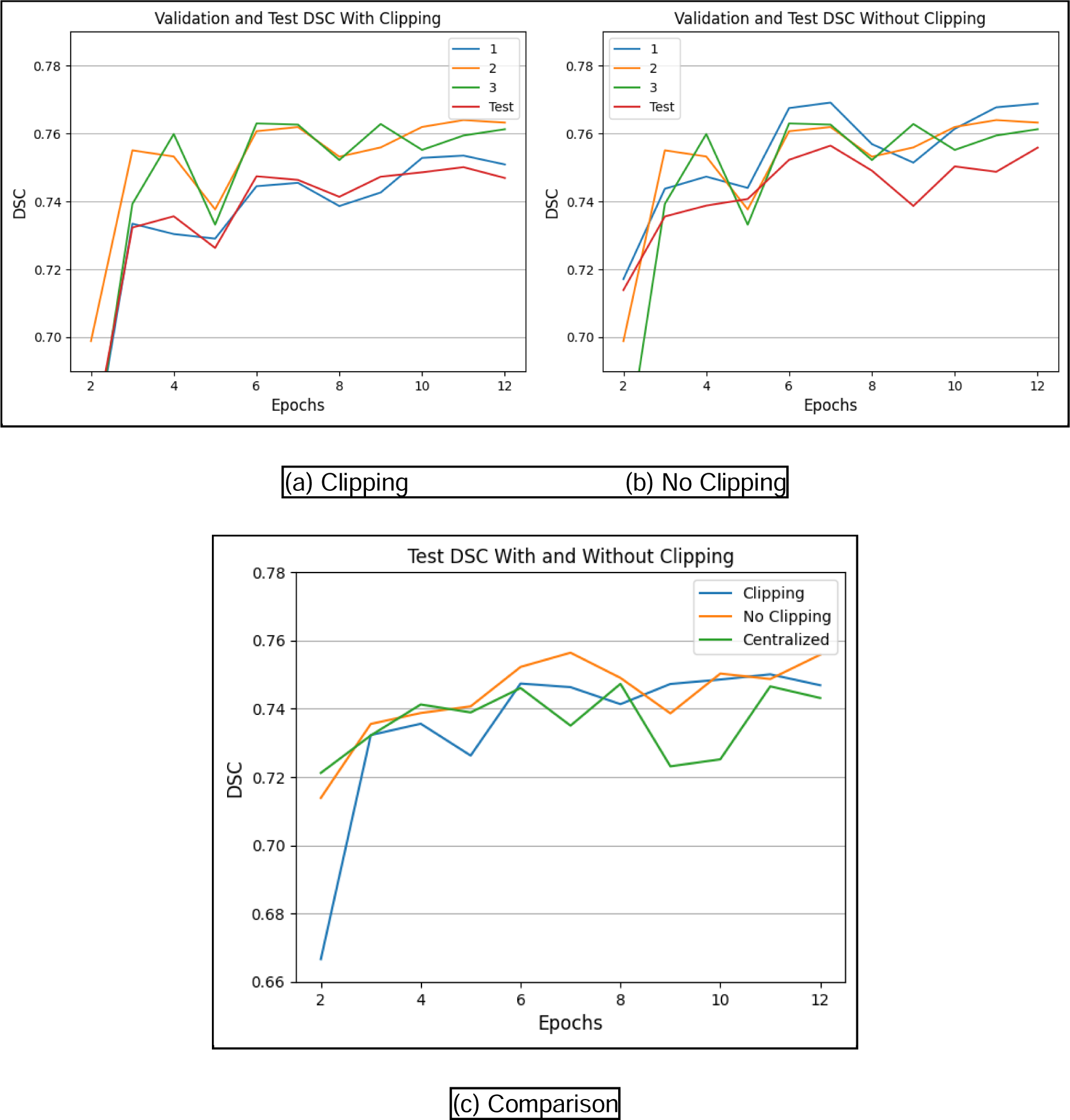
Task II Validation and Test DSCs. a) Validation DSC for clients 1-3 and Test DSC of the aggregated central model for rounds 2-12 with batch clipping. b) Validation DSC for clients 1-3 and Test DSC of the aggregated central model for rounds 2-12 without batch clipping. c) Comparison of the Test DSC for Clipping, No Clipping, and Centralized models. All Test DSCs in this figure were calculated using the first 2000 slices of the test set.

**Figure 5.**
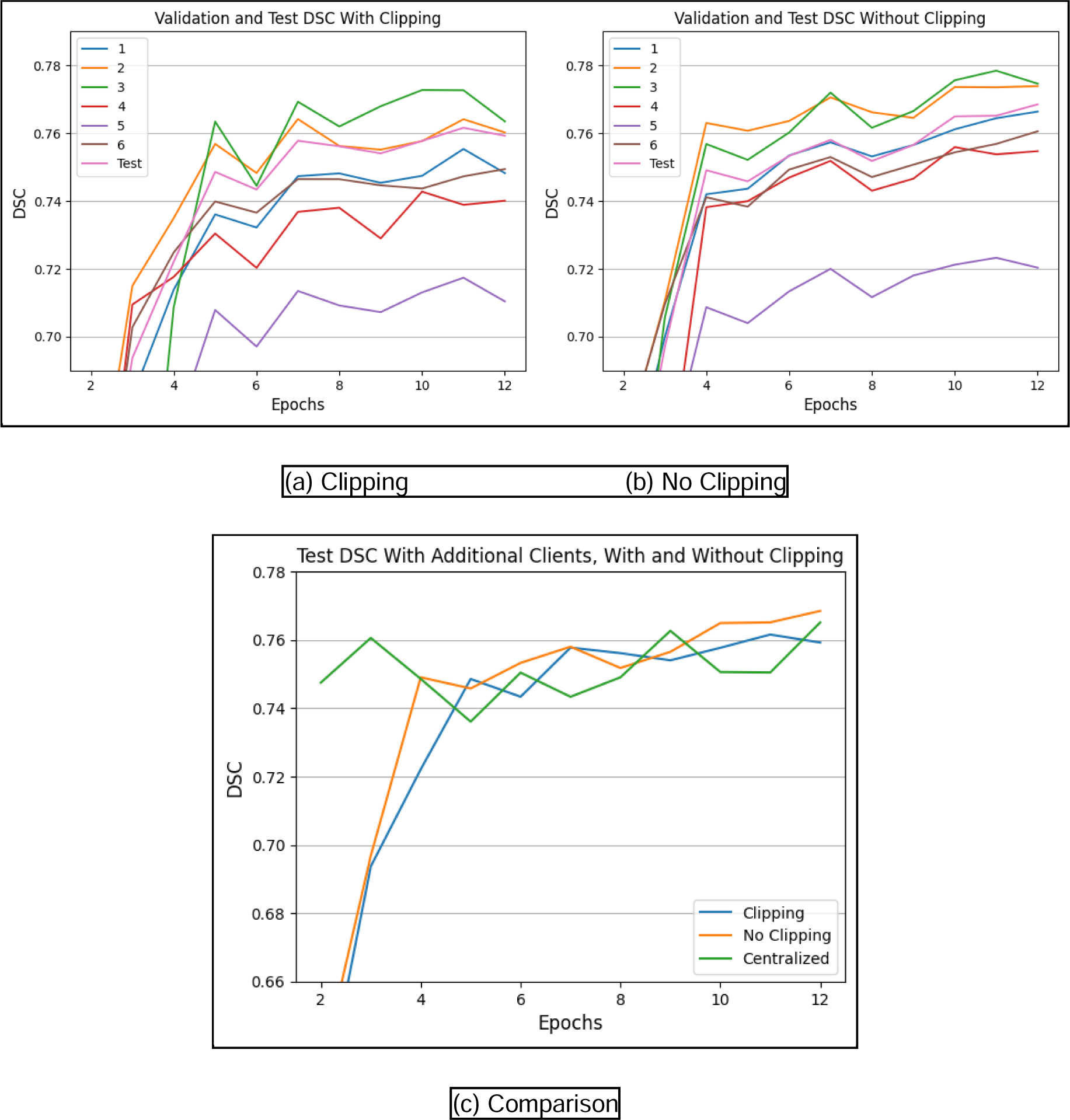
Task II Validation and Test DSCs. a) Validation DSC for clients 1-6 and Test DSC of the aggregated central model for rounds 2-12 with batch clipping. b) Validation DSC for clients 1-6 and Test DSC of the aggregated central model for rounds 2-12 without batch clipping. c) Comparison of the Test DSC for Clipping, No Clipping, and Centralized models when six clients are used. All Test DSCs were calculated using the first 2000 slices of the test set.

**Table 2.**
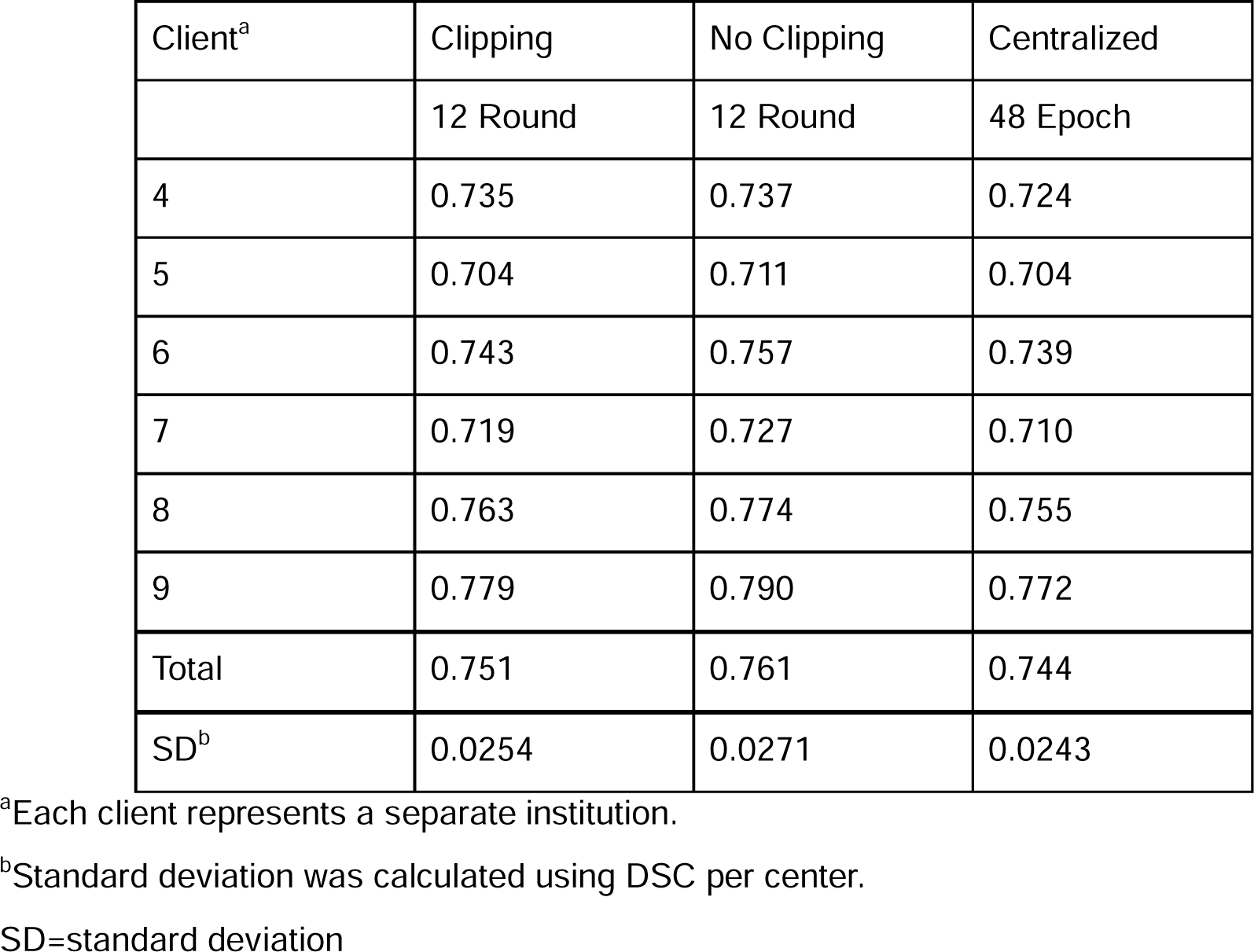
Task II average Dice similarity coefficient for each client in external test datasets.

### Additional Study: Task II training with Six Clients

In the additional study, we replicated the previous experiments of Task II with three additional clients, 4, 5, and 6, included in the training process (Figure S2). In the training dataset, client 5, the worst performing center in Task II trained on three clients, consistently exhibited lowest DSC even in the internal validation, scores 0.03∼0.05 less DSC than all other clients. Client 7 showed a large increase in DSC compared to the previous experiment, whereas DSCs of clients 8 and 9 does not change (Table S3). Again, a one-way ANOVA test of the average DSC of centers reveals that their differences are statistically insignificant (p=0.846).

## Discussion

This study compared the performance of FL and CL models, demonstrating the viability of FL in segmenting acute and chronic ischemic brain lesions. The variation in training epochs required to achieve convergence for the two tasks effectively represents the generalizability of FL across both lightweight and data-intensive tasks. In addition, we demonstrated that one-shot or few-shot communication approach yielded lower performance of FL models compared with constant communication. We also empirically demonstrated that batch clipping is a valid strategy for reducing training time without sacrificing robustness.

Given its partitioned nature, the BraTS dataset—a benchmark for brain image segmentation distributed across multiple centers—has been utilized in medical FL studies. Pati et al (3) trained on BraTS and other datasets to produce a state-of-the-art FL model. However, they did not start their training from a randomly initialized model but rather a “public initial model” pre-trained from the BraTS dataset. Li et al (17) also used BraTS to simulate the non-i.i.d nature of medical data and empirically shows multiple experiments for differential privacy. The scale (∼10,000 images) of our datasets surpasses that of the BraTS dataset, which adds value to the empirical results of our models’ performance.

Our findings suggest that FL necessitates constant communication between the clients and the central server, rather than relying solely on one or two rounds of aggregation after multiple epochs of individual training. Although a two-round aggregation for Task II yields reasonable performance, it lacks generalizability beyond each client’s training set, resulting in unsatisfactory final performance compared to FL models that communicate every four epochs. FedYogi and FedAvg struggle to effectively aggregate all parameters coherently in a single round, leading to poor performance after just a few rounds of aggregation.

Implementing batch clipping effectively reduced the training time by half compared to models without it. In FL, where training occurs in parallel across multiple institutions, the overall duration is dictated by the slowest client in each round rather than the total data size. Therefore, limiting the maximum number of batches trained for each epoch can reduce the total training time. Moreover, despite an increase in the number of clients in Task II for additional study, the training time remained relatively constant for FL, showcasing it’s the parallelized nature. In contrast, the training time of CL increased linearly with the size of the training data, consuming round 6.5 hours.

We believe that the efficacy of batch clipping fundamentally stemmed from the aggregation methods employed in our experiments—FedYogi and FedAvg. Both methods calculate the weighted average of updates, considering the number of batches processed by each client in every round. Thus, even though some clients train on less data per epoch, the norm of the total updated gradient remains balanced as updates from other clients are more heavily weighted. Although not extensively examined, we anticipate that batch clipping should be compatible with other aggregation algorithms such as KRUM (18) and median, as well as defensive mechanisms like Differential Privacy (19).

In Task I, the FL model performed better than the CL model in testing scenarios, which is not typical. In most studies, FL is considered to have a strict performance ceiling below CL (20). The expectation holds true in training and validation losses, where even though some clients score lower training and validation losses, the average FL loss is consistently greater than the CL loss. However, the potential reason behind the reversal of performance in the test set needs to be addressed. First, it could be that the averaging nature of FL could cancel out erratic updates in opposite directions. Indeed, Karimireddy et al. (21) suggests “bucketing,” which involves averaging some client submissions before aggregating, to reduce variance between clients. However, their study primarily focused on improving FL rather than directly comparing FL and CL. The other possibility is that the data of smaller clients in the training dataset happened to represent the test data better than those the larger clients, and hence act as a “well-represented” dataset. In CL, the updates induced by small amount of well-represented data could be diluted by data from different institutions. Even though FL still takes a weighted average of the number of data for each client when aggregating, the small client can train solely with its well-represented data, which can steer the aggregated model towards higher test scores regardless of training and validation losses. Indeed, there were some correlations between the small, performant clients such as client E and G and the external test dataset such as lesion area and slice thickness, depicted in Figure S3 and S4.

We acknowledge several limitations of the study. Most notably, all experiments were conducted in simulated environments, as our motivation was to translate an existing CL model into an FL model. Therefore, all data were preprocessed identically and maintained as high-quality, pixel-level segmentation data. In practice, such ideal conditions may need to be relaxed (22). Additionally, we had the advantage of using a CL model to evaluate FL performance. However, confirming convergence can be challenging when applying FL to new tasks without existing models. Some minor assumptions were made during the design of FL models. For instance, the batch size is set to 4 instead of 12 for the original Task I CL model due to GPU memory constraints during simulating. Similarly, the choice to set four epochs per round was arbitrary. We believe these assumptions have minimal impact on the major findings of our experiments; however, they are reported for potential investigations.

In conclusion, our study provides two FL brain lesion segmentation models that rival CL models, empirically supporting the suitability of FL in medical image segmentation. Through our experimentations, we used batch clipping for efficient training. Our findings may facilitate future studies involving a larger set of clients and more diverse datasets.

## Data Availability

Data generated or analyzed during the study are available from the corresponding author by request.

## Notes

### Competing Interest Statement

Hoyeon Lee, Jonghyeok Park, Myungjae Lee, and Wi-Sun Ryu are employees of JLK Inc., Seoul, Republic of Korea. Other authors had nothing to declare.

### Funding Statement

This study was supported by the Multiministry Grant for Medical Device Development (KMDF_PR_20200901_0098).

### Author Declarations

The study protocol was approved by institutional review board of Dongguk University Ilsan Hospital (2017-09-017).

