## Supplementary Text for "In-Silo Federated Learning vs. Centralized Learning for Segmenting Acute and Chronic Ischemic Brain Lesions"

### **Supplemental Materials**

#### Training Information

##### Task I

Batch Size = 4 (12 in CL)

Number of clients = 9

Optimizer = AdamW(betas=(0.9, 0.999), decay=1e-2, eps=1e-8, lr=5e=4)

Scheduler = LinearWarmupCosineAnnealingLR(warmup=10)

Aggregation = FedYogi(eta=1e-2, beta1=0.9, beta2=0.99, tau=1e-3)

Loss function = (2*BinaryCrossEntropyLoss+DiceLoss)/3

##### Task II

Batch Size = 8

Number of clients = 3 (6 in Additional Study)

Optimizer = Adam(betas=(0.9, 0.999), decay=1e-3/300, eps=1e-7, lr=1e-3)

Loss function = DiceLoss

#### Brain Image Analysis

**Acute ischemic lesion dataset** From May 2011 to November 2013, we consecutively enrolled 14,740 patients with ischemic stroke or transient ischemic attack admitted to 10 participating centers within seven days from symptom onset (Figure 1). After excluding 1,194 patients using the following criteria: contraindication to MRI (n=688), poor quality or unavailability of brain diffusion-weighted imaging (DWI) (n=321), and MRI registration error (n=185), leaving 13,546 patients amongst 9 institutions for training and validation and 1 institution for external testing. Previous study for CL utilized a total of 13,597 patients originated from 10 institutions. Brain DWIs were obtained using 1.5-Tesla (n=9,084) or 3.0-Tesla (n=2,923) MRI systems. DWI protocols were: b-values of 0 and 1,000 s/mm^2^, TR of 2,400–9,000 ms, TE of 50–99 ms, voxel size of 1×1×3–5 mm^3^, interslice gap of 0–2 mm, and thickness of 3–7 mm. Ischemic lesions on DWIs were segmented by six experienced researchers using an in-house software under the close guidance of an experienced vascular neurologist.

**Chronic ischemic lesion dataset** From May 2011 to November 2013, we consecutively enrolled 10,423 patients admitted to 9 participating centers within seven days from symptom onset. We excluded the following patients: contraindication to MRI (n=315), poor quality or unavailability of brain diffusion-weighted or fluid-attenuated inversion recovery (FLAIR) MRI (n=1,632), and MRI registration error (n=55), leaving 8,421 patients amongst 3 institutions for training and 6 institutions for testing. All data were forwarded from the previous CL study. Brain FLAIR MRI was performed on 1.5 Tesla (n=6,583) or 3.0 Tesla (n=1,803) MRI systems. FLAIR image protocols were TE 76–187 ms, TR 6000–11568 ms, voxel size 1×1×3–7mm^3^, spacing 0.3–1.0mm, slice thickness 3–7mm, FOV 175–280mm, and matrix size (row) 256–768. As previously reported, each patient's high signal intensity white matter lesions on FLAIR were manually segmented by six experienced researchers under careful supervision by an experienced vascular neurologist. When chronic white matter lesions on FLAIR and acute infarct lesions on DWI overlapped or were adjacent, we determined the extent and distribution of FLAIR WMH based on lesions in the hemisphere contralateral to the acute infarct location, as WMH symmetry of morphology and distribution are often observed between hemispheres.

#### Image Preprocessing

**Acute ischemic lesion dataset** Brain DWIs were preprocessed by applying Otsu's threshold and N4 bias correction (1,2). We adjusted DWI signals to fit between 0 and 255 before applying z-score normalization. Each image has been augmented with a linear combination of slice-wise similarity transform, MRI bias field simulation, axis flip, and gamma/contrast change.

**Chronic ischemic lesion dataset** We applied 2D B-spline interpolation to resize the FLAIR slices to a dimension of 256×256 pixels. Next, we performed slice-wise intensity normalization in a uint8 format to ensure pixel intensities ranged from 0 to 255 on each slice. We subsequently performed case-wise intensity equalization to ensure comparability in the imaging data across different MRI vendors by employing a histogram of 32 bins and shifting the highest peak of the histogram value to 150. Then, we generated a binary brain mask by including all voxels with signal intensities greater than a threshold value of 30 in the histogram domain. We filled tiny holes in this brain mask using binary_fill_holes from scipy.ndimage. Finally, we performed the Gaussian normalization process to yield pixel intensities with a mean of 0 and a standard deviation of 1 for the brain area under the brain mask, producing optimal arrays for training a deep learning model.
