## Supplementary Figure 1 for "In-Silo Federated Learning vs. Centralized Learning for Segmenting Acute and Chronic Ischemic Brain Lesions"

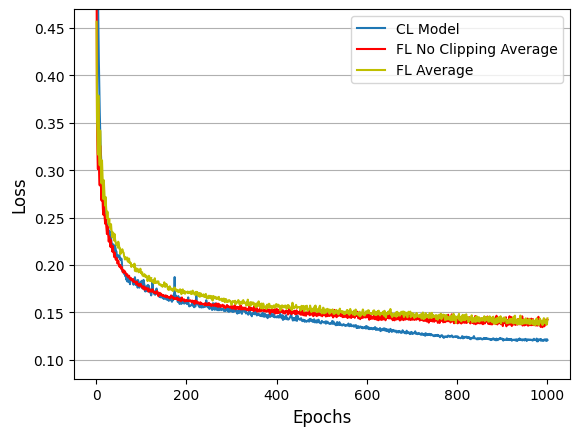

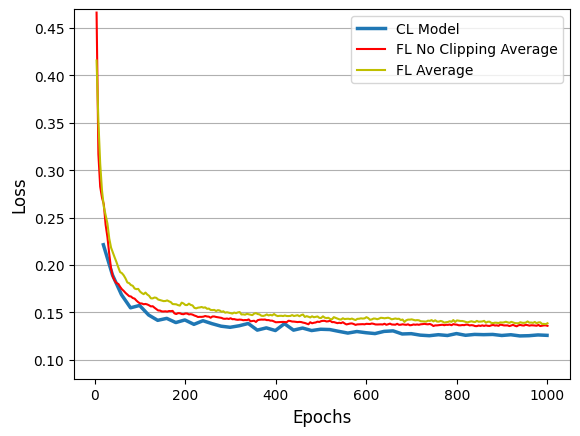

(a) Training Loss	(b) Validation Loss

**Figure S1. Losses and DSCs of CL and FL with and without Batch Clipping.** (a), (b) In both training and validation, FL without clipping performs on par with CL in the beginning but becomes similar to FL with clipping towards the end of training.
