## Supplementary Figure 2 for "In-Silo Federated Learning vs. Centralized Learning for Segmenting Acute and Chronic Ischemic Brain Lesions"

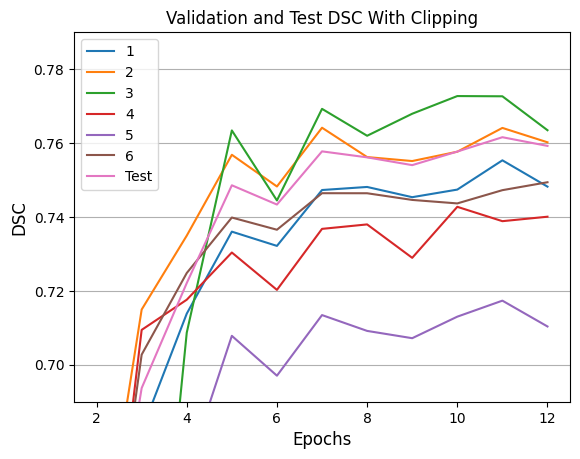

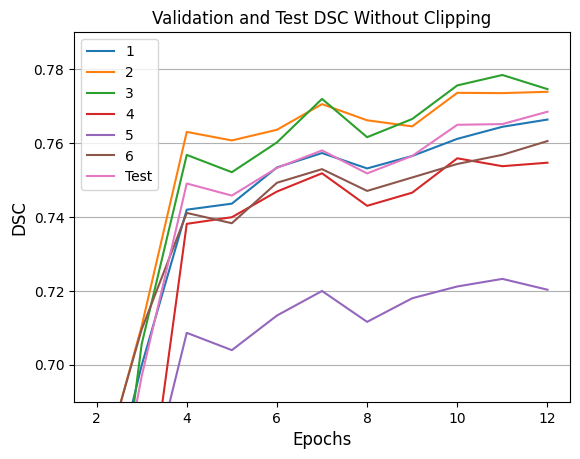

(a) Clipping	(b) No Clipping

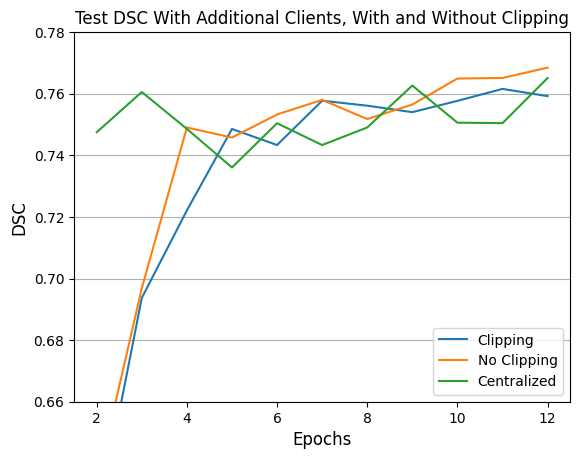

(c) Comparison

**Figure S2. Task II trained on six clients validation and test DSCs.** a) Validation DSC for clients 1-6 and Test DSC of the aggregated central model for rounds 2-12 with batch clipping. b) Validation DSC for clients 1-6 and Test DSC of the aggregated central model for rounds 2-12 without batch clipping. c) Comparison of the Test DSC for Clipping, No Clipping, and Centralized models when six clients are used. All Test DSCs were calculated using the first 2000 slices of the test set.
