## Supplementary Figure 3 for "In-Silo Federated Learning vs. Centralized Learning for Segmenting Acute and Chronic Ischemic Brain Lesions"

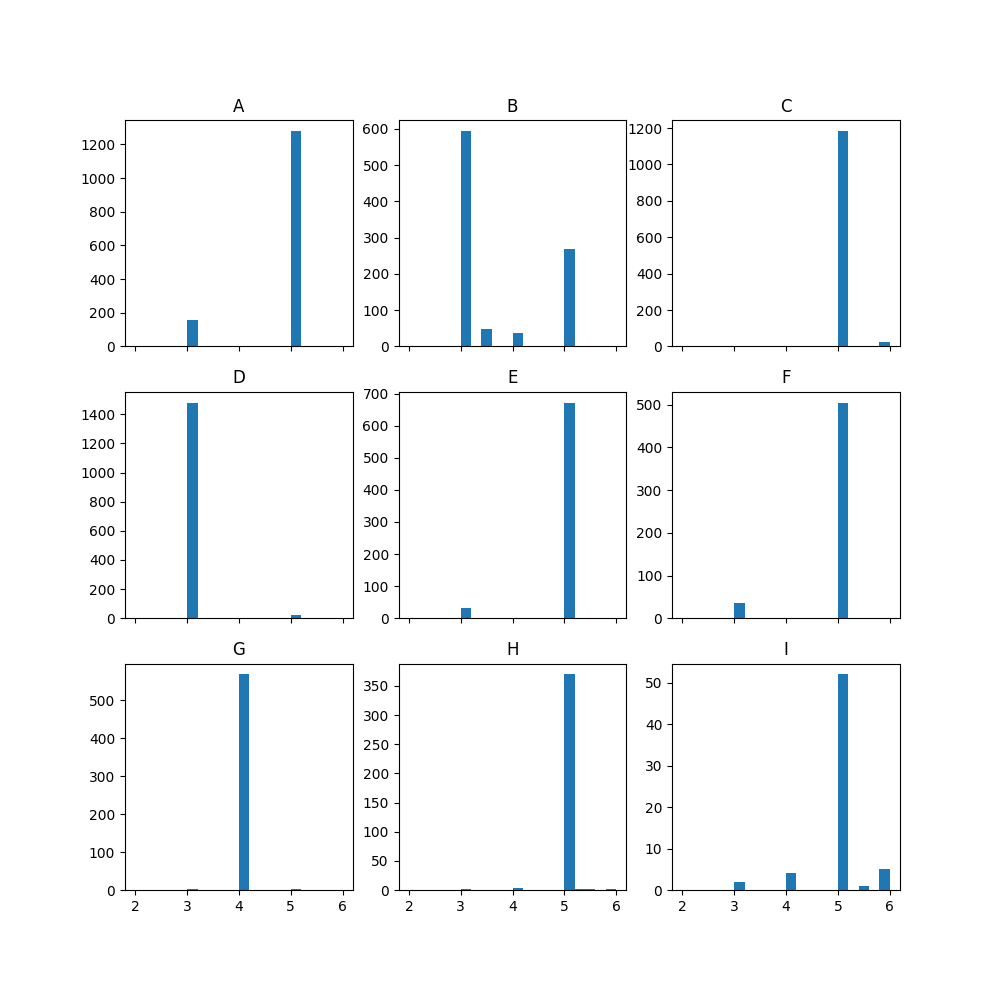

(a) Clients A-I in training dataset

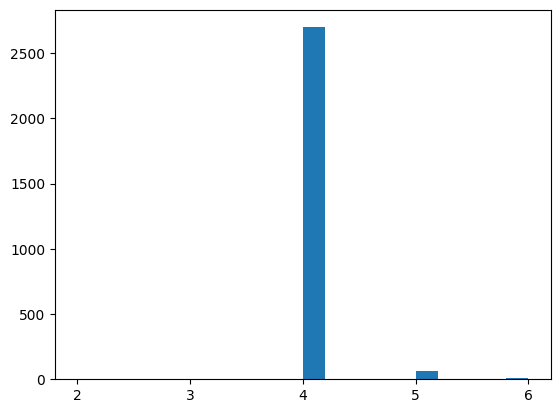

(b) External test dataset

**Figure S3. Slice thickness distributions for Task I.** (a) Histogram of slice thickness of images for clients A-I. Images without slice thickness information were excluded from counting. The y-axis of each subgraph is adjusted to fit the values of each client. (b) Histogram of slice thickness of images from the test set. The external test dataset primarily consisted of images of slice thickness of 4 mm. Of all clients, only client G shares this attribute, which is also the best performing client in FL training.
