## Supplementary Figure 4 for "In-Silo Federated Learning vs. Centralized Learning for Segmenting Acute and Chronic Ischemic Brain Lesions"

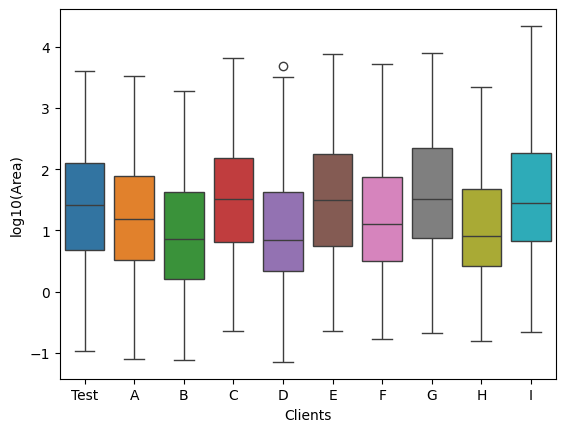

**Figure S4. Boxplot of logarithm of lesion volume for Task I.** Clients C, E, G, and I have a larger lesion area, while clients A, B, D, F, and H have a smaller lesion area.
