## Supplementary Table 1 for "In-Silo Federated Learning vs. Centralized Learning for Segmenting Acute and Chronic Ischemic Brain Lesions"

**Table S1.  Baseline characteristics of study population for Task I**

| Variables | Training & Validation  (n=10,769) | External test dataset  (n = 2,777) | P |
| --- | --- | --- | --- |
| Age (year)^a^ | 68.2 ± 12.7 | 68.2 ± 12.4 | .52 |
| Male^a^ | 5,546 (51.5%) | 1,571 (58.0%) | < .001 |
| Time from LKW to imaging^b^, median (IQR, hour) | 20.21 (5.2-49.5) | 11.41 (4.0–35.9) | < .001^c^ |
| Infarct volume, median (IQR, mL) | 1.91 (0.5-11.1) | 4.19 (0.8–19.4) | < .001^c^ |
| MRI vendor^d^ |  |  | < .001 |
| Phillips | 4,353 (40.4%) | 3 (0.1%) |  |
| GE | 2,146 (19.9%) | 2,706 (97.4%) |  |
| Siemens | 4,143 (38.5%) | 60 (2.2%) |  |
| Others | 9 (0.1%) | 8 (0.3%) |  |
| Magnetic field strength^e^ |  |  | < .001 |
| 1.5T | 6,309 (58.6%) | 2,724 (98.5%) |  |
| 3.0T | 2,882 (26.8%) | 41 (1.5%) |  |
| Pixel spacing (mm)^f^ |  |  | < .001 |
| < 0.8 | 1,646 (15.3%) | 11 (0.4%) |  |
| 0.8–0.849 | 1,732 (16.1%) | 11 (0.4%) |  |
| 0.85–0.899 | 2,675 (24.8%) | 10 (0.4%) |  |
| 0.9–0.949 | 1,329 (12.3%) | 12 (0.4%) |  |
| 0.95–0.999 | 652 (6.1%) | 55 (2.0%) |  |
| ≥ 1.0 | 2,735 (25.4%) | 2,676 (96.4%) |  |
| Slice thickness (mm)^g^ |  |  | < .001 |
| 3.0–3.9 | 2,908 (27.0%) | 1 (0.0%) |  |
| 4.0–4.9 | 781 (7.3%) | 2,699 (97.3%) |  |
| 5.0–5.9 | 5,475 (50.8%) | 66 (2.4%) |  |
| ≥ 6.0 | 36 (0.3%) | 8 (0.3%) |  |

Data are presented as mean ± standard deviation, number (percentage), or median (interquartile range, IQR). LKW=last-known-well.

^a^Data of age and sex were missing for 405 and 67 patients in the Training-and-validation dataset and the External dataset, respectively.

^b^Data of LKW to imaging time were missing for 5,447 and 1,849 patients in the Training & Validation dataset and the External dataset, respectively.

^c^Kruskal-Wallis test was used.

^d^Data of MRI vendors were missing for 118 in the Training & Validation dataset.

^e^Data of magnetic field strength were missing for 1,578 and 12 patients in the Training-and-validation dataset and the External dataset, respectively.

^f^Data of pixel spacing were missing for 2 patients in the External dataset.

^g^Data of slice thickness were missing for 1,569 and 3 patients in the Training-and-validation dataset, the Internal test dataset, and the External dataset, respectively.
