## Supplementary Table 2 for "In-Silo Federated Learning vs. Centralized Learning for Segmenting Acute and Chronic Ischemic Brain Lesions"

**Table S2.  Baseline characteristics of study population for Task II**

|  | Training & validation  (n=2,408) | External test 1  (n=1,105) | External test 2  (n=838) | External test 3  (n=2,654) | External test 4  (n=428) | External test 5  (n=571) | External test 6  (n=417) | *P* |
| --- | --- | --- | --- | --- | --- | --- | --- | --- |
| Age, year | 67.4±13.0 | 68.2±12.8 | 67.5±13.3 | 68.1±12.5 | 70.0±11.7 | 67.9±13.1 | 69.7±12.7 | <0.001 |
| Sex, men | 1,419 (60.4) | 568 (54.3) | 491 (60.8) | 1,497 (57.7) | 222 (52.7) | 346 (63.0) | 235 (58.6) |  |
| LKW to admission, hour | 12.0 (3.5-37.2) | 13.1 (3.5-32.0) | 13.9 (3.2-39.0) | 6.8 (2.6-24.0) | 11.4 (3.1-34.8) | 13.5 (3.3-39.4) | 17.7 (5.7-51) | <0.001^a^ |
| Revascularization | 404 (17.2) | 147 (14.1) | 82 (10.2) | 587 (22.6) | 73 (17.3) | 114 (20.8) | 45 (11.2) | <0.001 |
| Infarct volume, mL | 1.7 (0.4-9.8) | 1.1 (0.3-6.6) | 2.0 (0.4-12.2) | 3.6 (0.6-17.9) | 1.2 (0.3-8.7) | 2.2 (0.5-15.4) | 1.4 (0.5-9.5) |  |
| WMH volume, mL | 11.4 (5.4-24.7) | 14.6 (6.6-30.0) | 10.8 (4.2-25.1) | 12.7 (6.9-24.3) | 14.3 (7.0-28.7) | 13.0 (6.7-25.5) | 17.7 (8.9-40.0) | <0.001^a^ |
| **MRI vendor** |  |  |  |  |  |  |  | < 0.001 |
| Phillips | 1,027 (42.7%) | 1,098 (99.4%) | 0 | 0 | 0 | 1 (0.2%) | 299 (71.7%) |  |
| GE | 943 (39.2%) | 7 (0.6%) | 0 | 1,490 (56.1%) | 361 (84.4%) | 0 | 2 (0.5%) |  |
| Siemens | 420 (17.4%) | 0 | 0 | 12 (0.5%) | 67 (15.7%) | 569 (99.7%) | 116 (27.8%) |  |
| Others | 18 (0.8%) | 0 | 838 (100%) | 1,151 (43.4%) | 0 | 1 (0.2%) | 0 |  |
| **Magnetic field strength**^b^ |  |  |  |  |  |  |  | < 0.001 |
| 1.5T | 1,627 (67.6%) | 842 (76.2%) | 439 (52.4%) | 2,622 (98.8%) | 363 (84.8%) | 570 (99.8%) | 120 (28.8%) |  |
| 3.0T | 774 (32.1%) | 271 (23.6%) | 399 (47.6%) | 6 (0.2%) | 65 (15.2%) | 1 (0.2%) | 297 (71.2%) |  |
| **Acquisition matrix (row)** |  |  |  |  |  |  |  | < 0.001 |
| 256~511 | 421 (17.5%) | 55 (5.0%) | 3 (0.4%) | 35 (1.3%) | 427 (99.8%) | 570 (99.8%) | 0 |  |
| 512 | 1,972 (81.9%) | 1,019 (92.2%) | 455 (54.3%) | 2,612 (98.4%) | 1 (0.2%) | 1 (0.2%) | 366 (87.8%) |  |
| 513~768 | 15 (0.6%) | 31 (2.8%) | 380 (45.4%) | 7 (0.3%) | 0 | 0 | 51 (12.2%) |  |
| **Pixel spacing (mm)** |  |  |  |  |  |  |  | < 0.001 |
| 0.29 ~ 0.43 | 1,236 (51.4%) | 1,048 (94.8%) | 451 (53.8%) | 1,642 (61.9%) | 0 | 0 | 380 (91.1%) |  |
| 0.44 ~ 0.50 | 753 (31.3%) | 7 (0.6%) | 385 (45.9%) | 972 (36.6%) | 1 (0.2%) | 1 (0.2%) | 37 (8.9%) |  |
| 0.51 ~ 1.00 | 418 (17.4%) | 50 (4.5%) | 2 (0.2%) | 40 (1.5%) | 427 (99.8%) | 570 (99.8%) | 0 |  |
| **Slice thickness (mm)**^c^ |  |  |  |  |  |  |  | < 0.001 |
| 5 | 2,364 (98.2%) | 1,104 (99.9%) | 837 (99.9%) | 2,633 (99.2%) | 428 (100%) | 570 (99.8%) | 417 (100%) |  |
| **Repetition time (ms)** |  |  |  |  |  |  |  | < 0.001 |
| 6000 ~ 8499 | 1,076 (44.7%) | 156 (14.1%) | 0 | 73 (2.8%) | 0 | 3 (0.5%) | 291 (69.8%) |  |
| 8500 ~ 9999 | 368 (15.3%) | 949 (85.9%) | 397 (47.4%) | 2,574 (97.0%) | 66 (15.5%) | 567 (99.3%) | 118 (28.3%) |  |
| 10000 ~ 11568 | 964 (40.0%) | 0 | 441 (52.6%) | 7 (0.3%) | 361 (84.5%) | 1 (0.2%) | 8 (1.9%) |  |
| **Echo time (ms)** |  |  |  |  |  |  |  | < 0.001 |
| 75 ~ 119 | 425 (17.7%) | 331 (30.0%) | 0 | 30 (1.1%) | 67 (15.7%) | 570 (99.8%) | 116 (27.8%) |  |
| 120 ~ 143 | 1,677 (69.6%) | 768 (69.5%) | 837 (99.9%) | 463 (17.5%) | 0 | 1 (0.2%) | 300 (71.9%) |  |
| 144 ~ 187 | 306 (12.7%) | 6 (0.5%) | 1 (0.1%) | 2,161 (81.4%) | 361 (84.4%) | 0 | 1 (0.2%) |  |
| **Field of view (row, mm)** |  |  |  |  |  |  |  | < 0.001 |
| 175 ~ 219 | 654 (27.2%) | 819 (74.1%) | 2 (0.2%) | 26 (1.0%) | 409 (95.6%) | 9 (1.6%) | 28 (6.7%) |  |
| 220 | 974 (40.5%) | 279 (25.3%) | 70 (8.4%) | 1,638 (61.7%) | 5 (1.2%) | 480 (84.1%) | 340 (81.5%) |  |
| 221 ~ 280 | 780 (32.4%) | 7 (0.6%) | 766 (91.4%) | 990 (37.3%) | 14 (3.3%) | 82 (14.4%) | 49 (11.8%) |  |

Data are presented as mean±SD, median (interquartile range), number (percentage).

^a^Kruskal-Wallis test was used.

^b^Data were missing in a patient in external validation 3 dataset.

^c^Data were missing in 7, 2, and 26 patients in training and internal validation, external validation 1, and external validation 3 datasets, respectively.

LKW=last known well; WMH=white matter hyperintensity
