## Supplementary Table 3 for "In-Silo Federated Learning vs. Centralized Learning for Segmenting Acute and Chronic Ischemic Brain Lesions"

**Table S3. Task II average DSC for each client in the external test datasets after trained on 6 clients**

| Center | Clipping | No Clipping | Centralized |
| --- | --- | --- | --- |
|  | 12 Round | 12 Round | 48 Epoch |
| 7 | 0.739 | 0.741 | 0.731 |
| 8 | 0.760 | 0.777 | 0.773 |
| 9 | 0.778 | 0.793 | 0.781 |
| Total | 0.765 | 0.777 | 0.767 |
| SD | 0.0157 | 0.0218 | 0.0220 |

^a^Each client represents a separate institution.

^b^Standard deviation was calculated using DSC per center.

SD=standard deviation
